# Justifying model complexity: evaluating transfer learning against classical models for intraoperative nociception monitoring under anesthesia

**DOI:** 10.1101/2025.07.01.25330670

**Authors:** Chanseo Lee, Jaihyoung Lee, Kimon-Aristotelis Vogt, Muhammad Munshi

## Abstract

**Background:** Accurate intraoperative detection of nociceptive events is essential for optimizing analgesic administration and improving postoperative outcomes. While deep learning models promise to capture complex temporal dynamics of physiological signals, their added complexity may not always yield clinically meaningful gains compared to well-engineered classical approaches.

**Methods:** We evaluated two classical supervised models—L1-regularized logistic regression and Random Forests (with and without drug dosing features)—against a Temporal Convolutional Network (TCN) transfer-learning framework. We used a dataset of 101 adult surgical cases (~50,000 annotated nociceptive events over ~18,500 minutes) sourced from PhysioNet that tracked 30 physiologic and 18 drug-related features in 5-second windows. All models were assessed under a leave-one-surgery-out cross-validation, with AUROC and AUPRC as primary metrics. We further examined probability calibration (Platt scaling, isotonic regression) and four ensemble strategies—including a meta-learner, MLP, and a feature-conditioned gated network—to quantify the benefit of deep personalization.

**Results:** Drug-aware Random Forests achieved the highest discrimination (AUROC 0.716; AUPRC 0.399), significantly outperforming the TCN transfer-learning model (AUROC 0.649; AUPRC 0.311). Isotonic calibration reduced expected calibration error by over 80% but did not alter discrimination. None of the ensemble methods surpassed the standalone Random Forest, and the gated network consistently assigned > 84% weight to the classical model. Permutation importances revealed critical mechanistic features related to sympathetic physiologic response.

**Conclusions:** In this head-to-head benchmark, interpretable classical models on expertly curated features matched or exceeded the performance of a complex deep learning approach, while offering superior computational efficiency and transparency. These findings underscore the importance of rigorous comparative evaluation before adopting high-complexity AI solutions in clinical practice.

**Data Availability Statement:** All data was sourced from Subramanian et al. on PhysioNet under data usage agreement and proper citations in the manuscript. All code and analysis can be provided upon reasonable request. The authors plan to upload their code on GitHub.

**Competing Interests Statement:** The authors declare no conflict of interests or financial stakes in this work.

**Funding Disclosures:** There is no funding to declare for this work.

## Introduction

Intraoperative detection of nociceptive events is critical for optimizing analgesic administration, as inadequate pain monitoring can lead to both acute postoperative complications and chronic pain syndromes.^1,2^ Because of the subjective nature of pain, accurate monitoring of nociception during anesthesia remains elusive, resulting in inadequate dosing of analgesics and poor pain control.

There are prior studies on applying machine learning to detect intraoperative nociceptive events to guide analgesic dosing, such as the NOL monitor. NOL has demonstrated substantial clinical utility, reducing the average pain of post-anesthesia care unit patients.^3^ However, the proprietary pipeline required for NOL utilizes specialized hardware, and the inner workings are poorly understood, making the system an an algorithmic “black box.”

Instead, many classical algorithms such as Random Forests and logistic regressions have been applied to physiologic pain assessment due to their robustness and interpretability.^4^ In fact, Random Forests and logistic regression classifiers have been implemented in many predictive tools, including discrimination of pain states from electrodermal activity (EDA) and cardiovascular indices.

Continued advances in deep learning promise richer modeling of temporal and multimodal signals. Multimodal CNNs integrating electroencephalogram, photoplethysmography, and electrocardiogram signals demonstrate improved nociception detection compared to single-sensor methods.^5^ However, these deep architectures often require significant compute resources, risk overfitting on small dataset sizes, and may offer marginal gains over well-engineered classical models.^6^ This raises the question of whether added architectural complexity of deep learning truly translates to clinical benefit.^7^

To address this question, our study explores transfer learning via TCNs, adapting a globally trained network to patient-specific data, potentially enhancing personalized pain detection on small datasets.^8^ Despite numerous studies on either classical or deep learning methods for pain detection, few have rigorously benchmarked these approaches head-to-head in a clinically realistic setting. Moreover, the field lacks quantitative assessment of when increased model complexity justifies its additional cost and interpretability trade-offs. We address this gap by implementing a Leave-One-Surgery-Out (LOSO) framework to compare classical models to a deep-learning model. We also implemented a gated ensemble network that combines these two approaches to dynamically weigh model contributions and reveal the true value of personalization and model complexity.

This study is especially relevant in the context of large language models (LLMs) and AI pipelines gaining traction in medical tasks, from diagnostics to documentation. The emergence of these models raise substantial concerns regarding transparency, ethical risk, and resource allocation.^9,10^ While emerging foundations like TabPFN show that transformer-based tabular models can excel in small-data regimes, their applicability to high-resolution time-series physiologic data remains both untested and computationally demanding.^11^

Our findings highlight that well-curated feature sets and interpretable classical models can match or exceed the performance of complex deep learning frameworks on nociception detection, while dramatically reducing computational burden and enhancing clinician trust. These findings are supported by several prior literature, although the subject remains hotspot for debate.^6,7,10,12^ Our study underscores the importance of evaluating model complexity against utility and efficiency as the medical community explores deep learning architectures and LLMs for healthcare problems.^13^

## Methods

### Data source and pre-processing

The dataset was sourced from PhysioNet.^14,15^ Subramanian et al. compiled a prospective archive of multi-sensor, continuous physiologic recordings (derived from ECG, EDA) and real-time drug dosing from 101 adult surgical cases, paired with manual annotations by anesthesiologists of 50,000 surgical nociceptive stimuli across ~18,500 minutes of surgery. 15 autonomic features and their respective estimated first derivatives for a total of 30 physiologic features, and 18 drug dosing chronology covariates (time since dose, cumulative dose) from nine drug classes.

The data was concatenated into a single-table with non-overlapping 5-second windows and then underwent quality assurance checks (eg. zero imputation for missing values in drug dosing). Every numeric column was then standard ⍰ scaled across the entire pooled cohort to zero mean and unit variance, ensuring equal weight during model fitting. For reproducibility, we created two input matrices: a 48⍰column version that includes both autonomic and drug covariates, and a 30 ⍰ column version that excludes the drug information. The manual annotated nociceptive stimuli recordings were used as the ground truth for comparison.

### Model creation and performance evaluation

Each model’s creation and performance were completed using a Leave-One-Surgery-Out (LOSO) cross-validation strategy. In this approach, data from each surgery was held out in turn as the test set, while the models were trained and saved on the remaining surgeries. This process was repeated for all surgeries, ensuring that each subject contributed exactly once as a test case.

For each LOSO fold, the held-out surgery was further partitioned for transfer learning experiments. For transfer-learning models, the initial segment of the surgery was used for patient-specific adaptation (fine-tuning), while the remainder was reserved for evaluation. The adaptation window was varied to assess the impact of patient-specific data on model performance.

Model discrimination was quantified using the Area Under the Receiver Operating Characteristic Curve (AUROC) and the Area Under the Precision-Recall Curve (AUPRC). 95% confidence intervals for the median AUROC and AUPRC were calculated using non-parametric bootstrapping with 10,000 resamples. To assess the statistical significance of differences between models, pairwise comparisons of AUROC and AUPRC distributions were performed using non-parametric Wilcoxon signed-rank test.

### Producing benchmark supervised models

To establish a performance benchmark, we created baseline models based on Subramanian et al.^1^ Four models were implemented: two logistic regression models with L1 regularization (LASSO), selected via the Akaike Information Criterion (AIC), with and without inclusion of pharmacologic features; and two random forest classifiers, each consisting of 200 decision trees with a maximum depth of 50, trained using 90% bootstrap resampling to mitigate overfitting. For ensemble experiments, an additional 50-tree random forest classifier with drug information was also trained with the same methodology.

### Transfer-learning models with adaptive windows

Each transfer-learning experiment was structured as a two-phase, leave-one-subject-out protocol. First, a global base model was initialized by pooling all 5-second windows from 100 of the 101 surgeries and training a lightweight Temporal Convolutional Network (TCN). This network applies a single 1-D convolution across the feature channels (48 channels when drug covariates are included, 30 otherwise), followed by batch-normalization, ReLU, global max-pooling, and a two-layer dense head. We optimized all parameters for up to twelve epochs (Adam, α = 1 × 10□^3^, batch = 128) with binary cross-entropy loss weighted for the 6% event prevalence, using early stopping (patience = 3) to avoid over-fitting. The resulting weights were checkpointed as the base model for that LOSO fold.

In the personalization phase, we loaded the base model, froze its convolutional and normalization layers, and fine-tuned only the dense head on the first K minutes of the held-out patient’s own data (where K ∈ {1, 2, 5, 10}, corresponding to 12, 24, 60, or 120 windows). Fine-tuning ran for three epochs (Adam, α = 1 × 10□ □, batch = 128) to prevent forgetting and optimizing for small dataset size. The resulting models were saved for each patient in a LOSO-fashion.

### Calibration analysis

To assess the reliability of predicted probabilities from the transfer learning models, calibration analysis was performed across adaptation window lengths. For each window, the TL model was evaluated on the held-out portion of each surgery in the LOSO cross-validation framework. Predicted probabilities and true labels were aggregated for each adaptation window.

Three calibration approaches were compared: raw (uncalibrated), in which direct probabilities are output from the TL model; Platt scaling, in which a logistic regression model was fit to map the raw outputs to calibrated probabilities; isotonic regression, in which a non-parametric isotonic regression model was fit to the raw outputs.

Calibration performance was assessed using reliability curves, the Brier score, and the expected calibration error (ECE), computed with 10 quantile-based bins. For each method, calibration curves were plotted by comparing the mean predicted probability to the observed event frequency within each bin.

### Ensemble methods

To further enhance predictive performance, several ensemble strategies were evaluated by combining the outputs of the RF and TL models. First, a simple linear combination was implemented, where the final prediction was a weighted average of the RF and TL model outputs, with weights either fixed or optimized via linear regression on the validation data. Additionally, a pruned version of the RF models (with 50 trees instead of 200) was employed to observe the behavior of the resulting ensemble models compared to the 200-tree baseline.

Meta-learning approaches were explored beyond linear combination. A one-layer meta-learner, implemented as logistic regression, was trained to learn optimal weights for combining the base model predictions. For greater flexibility, a two-layer neural network meta-learner was also evaluated, allowing the ensemble to capture potential non-linear relationships between the base model outputs.

Finally, a gated network (GateNet) ensemble was implemented. In this approach, a small neural network was trained to dynamically assign input-dependent weights to the RF and TL predictions, effectively learning when to rely more on one model versus the other based on the input features. All ensemble models were trained and evaluated within the same LOSO cross-validation framework as the base models, ensuring fair and unbiased comparison. Pooled AUROC/AUPRC as well as two-tailed Wilcoxon tests were between ensembles and base models were calculated.

## Results

### Personalization of nociceptive signal detection

**Table 1** delineates the model performances of the Random Forest and Logistic Regression baselines that are drug-naïve and drug-aware. The RF models consistently outperform LR in terms of AUROC and AUPRC with statistical significance. Interestingly, the RF models also benefit from intraoperative drug information (AUROC 0.716 [0.700, 0.759]) versus without (AUROC 0.662 [0.640, 0.700]) with statistical significance. However, this pattern is not reflected in the LR models.

**Table 1.** Classical models as a baseline for nociceptive signal discrimination.

|  | RF (aware) | RF (naïve) | LR (aware) | LR (naïve) |
| --- | --- | --- | --- | --- |
| **AUROC** | 0.716 | 0.662 | 0.623 | 0.624 |
| *95% CI* | [0.700, 0.759] | [0.640, 0.700] | [0.583, 0.643] | [0.602, 0.649] |
| **AUPRC** | 0.399 | 0.339 | 0.291 | 0.291 |
| *95% CI* | [0.370, 0.434] | [0.309, 0.364] | [0.265, 0.317] | [0.266, 0.324] |

Classical models and their reported median AUROC/AUPRC; RF: Random Forest; LR: Logistic Regression; aware models take 48 inputs including 18 drug features; naïve models take 30 physiologic inputs only; 95% confidence intervals reported in brackets.

The performance of transfer-learning models is shown in **Table 2**. At face value, the models’ AUROC/AUPRCs benefit marginally without statistical significance either from drug information or increased personalization phases despite being introduced up to 10 times more data. However, a granular per-surgery AUROC benefit analysis between 10 minutes and 1 minute of personalization **(Fig. 1)** showed that 67 of 101 surgeries (66%) improved from the additional adaptation in drug-aware models, while 34 (34%) declined. The median AUROC benefit was 0.019 with an interquartile range of [-0.023, 0.047].

**Table 2.** Drug-aware and naïve transfer-learning models with varying adaptation windows.

| **Adaptation**<br>**(Aware)** | **1 min.** | **2 min.** | **5 min.** | **10 min.** |
| --- | --- | --- | --- | --- |
| **AUROC** | 0.641 | 0.640 | 0.639 | 0.649 |
| *95% CI* | [0.616, 0.663] | [0.619, 0.659] | [0.618, 0.664] | [0.619, 0.669] |
| **AUPRC** | 0.317 | 0.326 | 0.301 | 0.311 |
| *95% CI* | [0.285, 0.358] | [0.301, 0.366] | [0.282, 0.365] | [0.284, 0.366] |
| **Adaptation**<br>**(Naïve)** | **1 min.** | **2 min.** | **5 min.** | **10 min.** |
| **AUROC** | 0.632 | 0.636 | 0.632 | 0.638 |
| *95% CI* | [0.611, 0.658] | [0.604, 0.651] | [0.612, 0.649] | [0.610, 0.665] |
| **AUPRC** | 0.314 | 0.316 | 0.309 | 0.323 |
| *95% CI* | [0.273, 0.356] | [0.274, 0.358] | [0.272, 0.343] | [0.288, 0.346] |

Drug-aware and drug-naïve transfer learning models with adaptation windows from 1-10 minutes; reported median AUROC, AUPRC; 95% confidence intervals reported in brackets.

**Figure 1.**
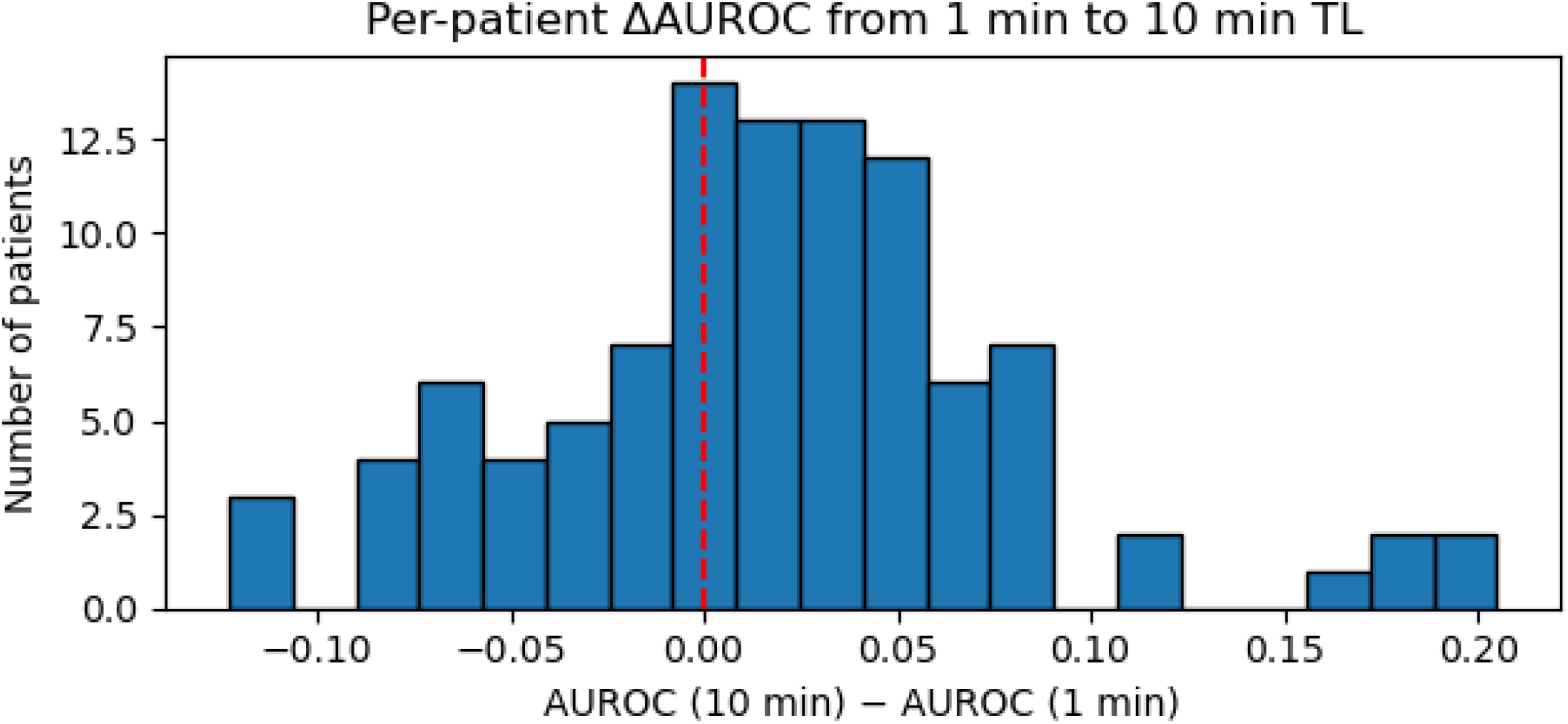
Per-patient AUROC change between 1-minute to 10-minute adaptation windows. AUROC differences were calculated for each personalized surgery (LOSO fold) then plotted in the histogram above. 66% of surgeries experienced an increase in AUROC while 34% experienced a decrease.

### Isotonic calibration improves model accuracy

Calibration analysis, at its simplest, is meant to show whether an **X** % predicted risk by the transfer learning model directly translates to **X** % of cases experiencing a nociceptive signal. We find that while the three calibration methods do not significantly improve its discriminatory ability in terms of AUROC **(Table 3)**, the Brier scores and ECE indeed show significant improvement from the raw model, approaching nearly zero for all adaptation windows **(Fig. 2A)**. Platt scaling also provided some improvement (ECE_10 min_ = 0.0198), but the reliability curves still deviated from the ideal diagonal **(Fig. 2B)**, especially in the region where fewer positive samples were available. A Wilcoxon two-tailed test showed that the isotonic calibration performed significantly better than both Platt scaling and the raw models (p < 0.001).

**Table 3.** Calibration of 10-minute drug-aware transfer learning models and skipped-folds analysis.

| Calibration | Uncalibrated | Platt | Isotonic |
| --- | --- | --- | --- |
| **AUROC** | 0.649 | 0.649 | 0.663 |
| *95% CI* | [0.619, 0.669] | [0.632, 0.671] | [0.643, 0.671] |
| **Brier** | 0.196 | 0.158 | 0.155 |
| *95% CI* | [0.184, 0.208] | [0.155, 0.171] | [0.151, 0.165] |
| **ECE** | 0.151 | 0.051 | $7.14 \times 10^{-9}$ |
| *95% CI* | [0.1318, 0.171] | [0.044, 0.056] | $[5.58 \times 10^{-9}, 9.42 \times 10^{-7}]$ |

| Adaptation Time | Skipped Folds (out of 101) |
| --- | --- |
| 1 min. | 96 |
| 2 min. | 89 |
| 5 min. | 48 |
| 10 min. | 8 |

Calibration analysis of transfer-learning models and their reported median AUROC, Brier, and Expected Calibration Error scores (ECE). Statistical significance achieved in ECE scores with paired Wilcoxon in isotonic vs. Platt ( $p = 0$ ) and isotonic vs. uncalibrated ( $p = 0$ ). Skipped folds analysis analyze whether the adaptation windows contained sufficient information to perform Platt calibration.

**Figure 2.**
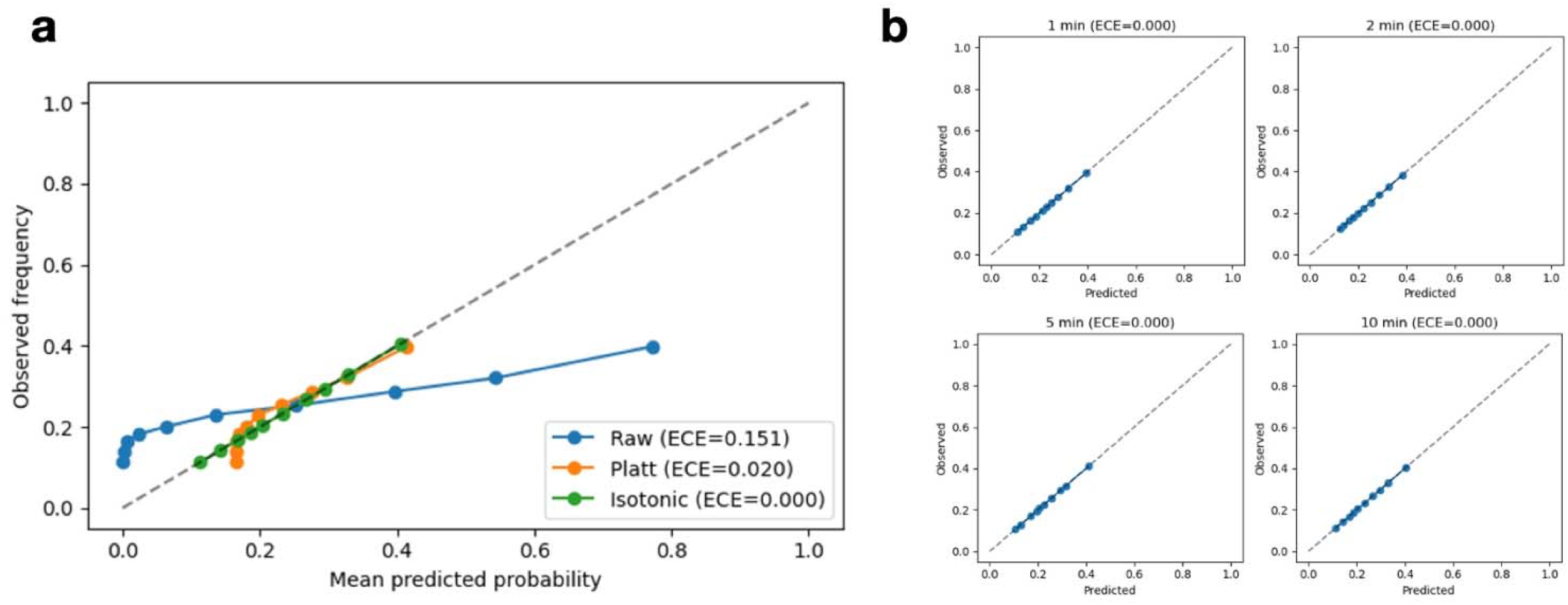
Calibration analysis of transfer-learning with varying adaptation windows **a**. Mean-predicted probability plotted against observed frequency for the three calibration strategies. The diagonal dashed line represents the ideal calibration where observed frequency equals the predicted probability. **b**. Isotonic calibration across 1-, 2-, 5-, and 10-minute adaptation windows and their respective Expected Calibration Error (ECE).

Across adaptation windows, the number of “skipped folds” **(Table 3)**—cases where Platt scaling could not be performed due to a lack of both positive and negative events in the adaptation set—decreased substantially as the adaptation window increased. For the shortest window (1 minute), Platt scaling was feasible in only 5 out of 101 cases, with 96 folds skipped. As the adaptation window lengthened, the number of skipped folds declined, reaching 8 at the 10-minute window.

### Ensemble modeling reveals the key insights into intraoperative nociception prediction

Our baseline Random Forest models and tailored transfer-learning models were ensembled together with the hypothesis that they could not only improve performance but could also reveal key insights about the operative data and best practices when applying deep learning to nociception. The four ensemble strategies include simple linear combination, one-layer logistic regression meta-learner, two-layer non-linear meta-learner, and dynamic GateNet.

An initial comparison between a linear combination ensemble of a 200-tree RF versus a pruned 50-tree RF with the 10-minute TL model revealed that while both the RF and RF-TL ensembles outperform the TL model, there was no statistical difference in performance with increasing RF tree number **(Table 4)**. The pruned RF(50)-TL (AUROC 0.681 [0.679, 0.684]) and unpruned RF(200)-TL (AUROC 0.683 [0.681, 0.686]) performed similarly to each other, but worse than the RF models alone. The pruned RF (AUROC 0.715 [0.713, 0.718]) and unpruned RF (AUROC 0.713 [0.711, 0.716]) also performed similarly to each other.

**Table 4.** Pooled AUROC and AUPRC comparison of RF-TL ensemble methods.

| Model | Base models |  |  | Simple linear combination |  |
| --- | --- | --- | --- | --- | --- |
|  | RF(50) | RF(200) | TL (10 min.) | RF(50)-TL | RF(200)-TL |
| **AUROC** | 0.715 | 0.713 | 0.635 | 0.681 | 0.683 |
| 95% CI | [0.713, 0.718] | [0.711, 0.716] | [0.632, 0.637] | [0.679, 0.684] | [0.681, 0.686] |
| **AUPRC** | 0.409 | 0.415 | 0.323 | 0.359 | 0.372 |
| 95% CI | [0.404, 0.413] | [0.410, 0.419] | [0.319, 0.326] | [0.355, 0.363] | [0.367, 0.376] |
| **ENS vs. RF** | **Not applicable** | | | $4.88 \times 10^{-6}$ | $2.43 \times 10^{-6}$ |
| **ENS vs. TL** | **Not applicable** | | | $8.08 \times 10^{-13}$ | $2.63 \times 10^{-13}$ |

| Model | One-layer meta-learner |  | Two-layer MLP | GateNet |
| --- | --- | --- | --- | --- |
|  | RF(50)-TL | RF(200)-TL | RF(200)-TL | RF(200)-TL |
| **AUROC** | 0.686 | 0.708 | 0.710 | 0.712 |
| 95% CI | [0.684, 0.688] | [0.706, 0.711] | [0.708, 0.713] | [0.710, 0.715] |
| **AUPRC** | 0.365 | 0.413 | 0.415 | 0.416 |
| 95% CI | [0.361, 0.369] | [0.409, 0.418] | [0.411, 0.420] | [0.412, 0.421] |
| **ENS vs. RF** | $2.53 \times 10^{-5}$ | $1.70 \times 10^{-5}$ | **0.221** | **0.580** |
| **ENS vs. TL** | $1.26 \times 10^{-12}$ | $1.09 \times 10^{-8}$ | $8.69 \times 10^{-11}$ | $1.65 \times 10^{-10}$ |

RF(X): Random Forest with X trees; TL: transfer-learning; RF-TL: ensemble between RF-TL; MLP: multi-layer perceptron used for ensemble; GateNet: feature-conditioned gated neural network used for ensemble; ENS vs. RF/TL: comparing ensemble performance to RF or TL using paired Wilcoxon tests on pooled AUROC. Above threshold $p > 0.05$ indicates no statistical difference between ensemble and base model performance.

To investigate the effects of ensemble behavior between RF and TL in nociception detection, a one-layer meta-learner was employed. Interestingly, while the pruned RF-TL (AUROC 0.686 [0.684, 0.688]) did not improve in performance, the unpruned RF-TL (AUROC 0.708 [0.706, 0.711]) performed significantly better than its pruned counterpart, despite the pruned RF(50) and unpruned RF(200) having no statistical difference. However, the unpruned RF-TL with the one-layer meta-learner still significantly underperformed than its unpruned RF counterpart (p < 0.001).

To test a further hypothesis that the interaction between model outputs may be non-linear, we utilized a two-layer meta-learner to potentially boost ensemble performance. The RF(200)-TL (AUROC 0.710 [0.708, 0.713]) performed similarly to the RF(200) (AUROC 0.713 [0.711, 0.716]) without significant difference (p = 0.221).

### Feature-conditioned gated neural networks allow ensemble interpretation

An additional ensemble strategy employing a feature-conditioned gated neural network was used to reveal mechanistic insights of nociceptive signal prediction. The GateNet performed as well as the Random Forest, achieving an AUROC of 0.712 [0.710, 0.715]. A similar per-surgery analysis revealed that 60 of 101 (59.4%) surgeries experienced an AUROC increase while 41 declined (40.6%). However, the median ΔAUROC was 0, and the IQR was [0, 0.001], suggesting spurious improvements.

Permutation importances were calculated from the Random Forest, RF(200)-TL ensemble, and the GateNet in **Figure 3**, which delineates the top 10 features from each calculation. We note that both the ensemble and Random Forest have high dependencies on time since last sedative dose, tonic electrodermal activity, and the mean heart rate, indicating a high dependency on these factors for predictive power and optimal weight assignments.

**Figure 3.**
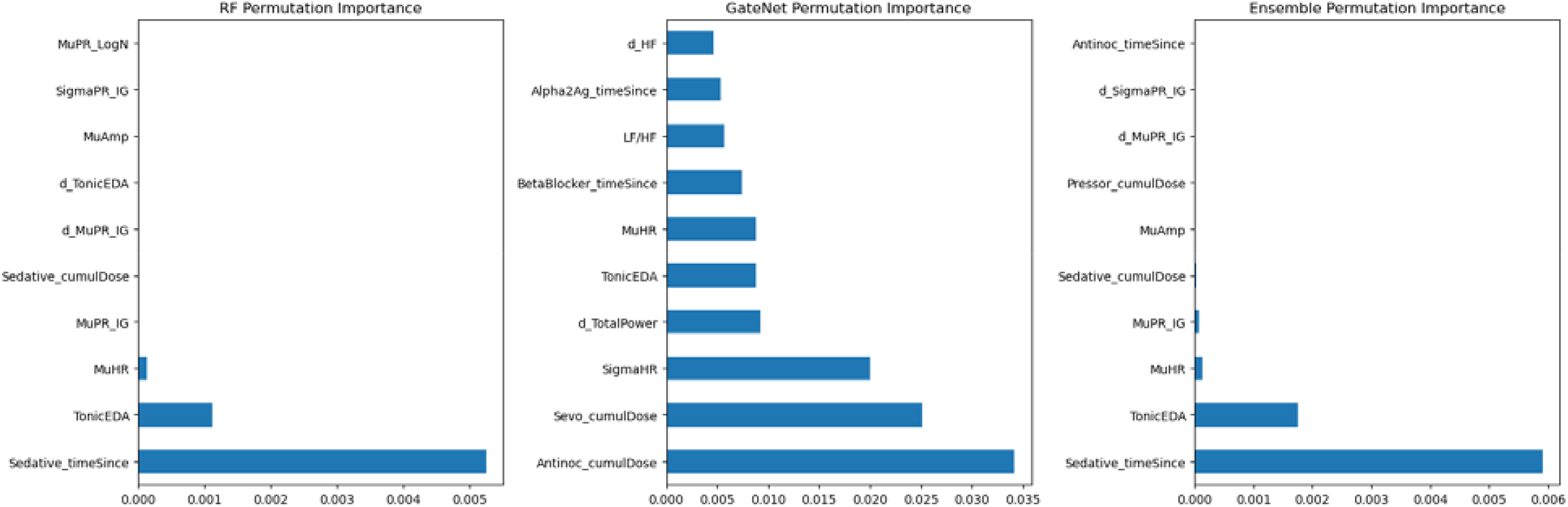
Permutation importance of predictive driver features in RF, ensemble, and GateNet. Top 10 permutation importance features for Random Forest (RF), GateNet arbitrator, and RF(200)-TL ensemble model. Note that “Mu-” stands for mean, “Sigma-” stands for standard deviation, and “d_-“ indicates first derivative features.

GateNet’s top three features included cumulative doses of antinociceptives, sevoflurane, and heart rate variability. Furthermore, unlike the Random Forest and the ensemble, it had proportionally similar, lesser important features such as Tonic EDA and time since beta blocker and alpha-2 agonist dosing. These features were important in determining levels of trust between the RF versus TL models.

Given the high permutation importances observed with the cumulative doses of antinociceptives and sevoflurane, the GateNet α was plotted over these two features. **Figure 4A** depicts α over observed evaluation windows, and **Figure 4B** shows a heatmap of α over all simulated evaluation windows when independently varying the two features while others are held constant. Both show that α always trends near 1 with a minimum of 0.84, indicating the ensemble relied more on RF than TL to make final prediction decisions.

**Figure 4.**
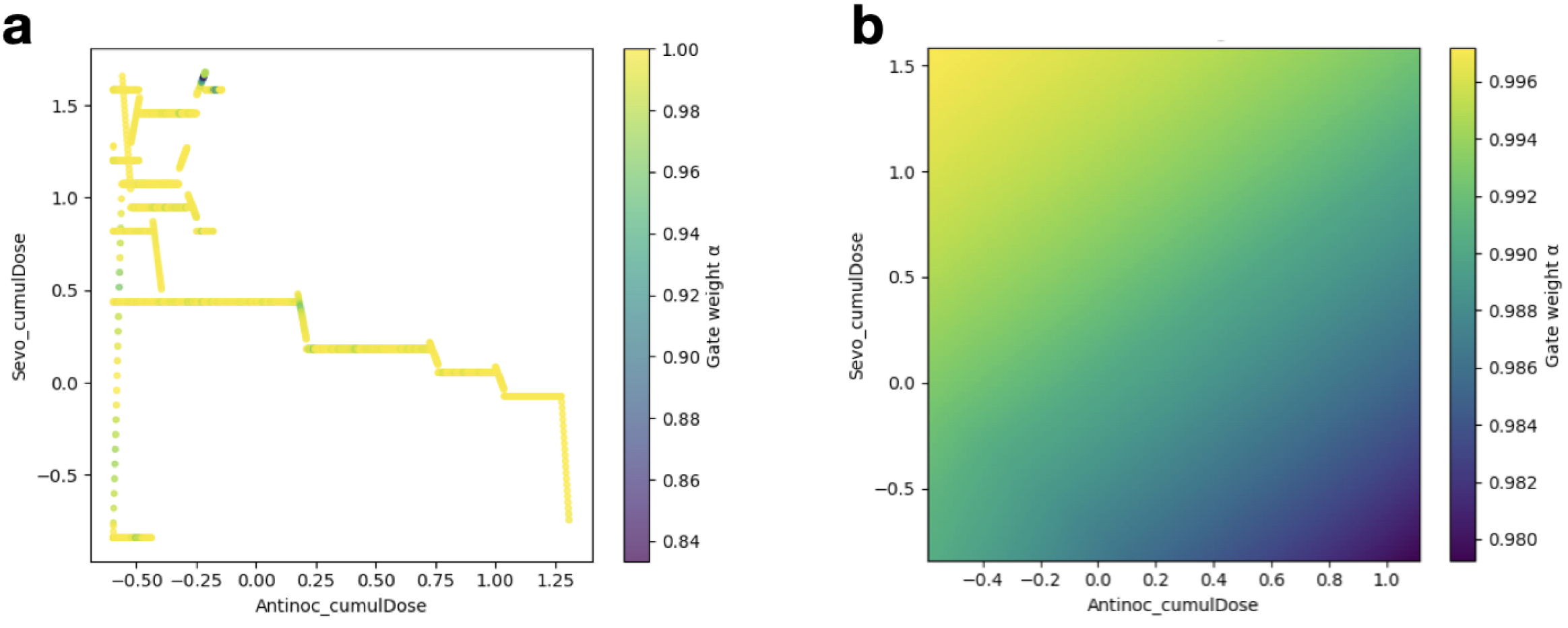
GateNet α analysis for across antinociceptive and sevoflurane dosing features. **a**. GateNet α plotted over observed evaluation windows. The colors and their corresponding α are shown in the color bar. **b**. Simulated α values across simulated evaluation windows by independently varying the respective features.

## Discussion

Accurate detection and management of pain during surgery remains a critical challenge in perioperative medicine, with significant implications for patient outcomes and the advancement of personalized care. Intraoperative nociception is inherently complex, influenced by a dynamic interplay of physiological responses and pharmacological interventions. Traditional monitoring approaches often fail to capture this complexity, leading to suboptimal pain control and increased risk of adverse events.

Prior literature has explored machine learning techniques to improve intraoperative pain assessment. However, questions surrounding the clinical utility of machine learning methodologies in the context of small data sizes and interpretability remain largely unanswered. Our study is the first to address these questions by introducing a flexible ensemble framework that integrates both classical and deep learning models, guided by a neural gating mechanism. This approach provides interpretable insights into the relative importance of physiological and drug-related features and analyzing methodologies for personalized pain detection. We harness ensemble methods and data science to assess the cost-benefit of increasing model complexity.

### Personalized deep learning doesn’t always outperform reliable supervised learning techniques

Our study found that architecture is not the only factor in creating clinically effective machine learning and artificial intelligence in precision medicine. Despite having the advantage of additional study of up to 10 minutes of a patient’s surgery in each LOSO fold, we find that transfer learning models do not outperform the Random Forest baseline. We provide several pieces of literature evidence supporting this finding.

First, the Random Forest is commonly used for performing predictions on medical tabular data^11^ because of its strong performance on irregular, non-rotationally invariant data where linear combinations of features may be uninformative for predictive power.^12^ In these cases, tree-based methods like Random Forest commonly outperform deep learning methods.^7,12^

Second, the models in this study were fed expertly crafted features, such as mean, standard deviations, first derivatives, drug timings, and dosages. These features may already capture high amounts of domain knowledge and pattern discovery.^16^ For example, permutation importance revealed factors such as TonicEDA, mean heart rate, or standard deviation of heart rate as high importance features in our ensemble. These features have been shown to be associated with sympathetic chain function and nociceptive stimuli and response.^17,18^

Against a 200-tree Random Forest trained on 100 diverse surgeries, the TCN-based transfer learning model, despite traditionally excelling on small sample sizes on prior medical detection studies,^19,20^ offers marginal benefits in a highly engineered feature set. In fact, fine-tuning on just the first few minutes of surgery could be detrimental, because this initial period can be noisy, not contain enough information, or not representative of the rest of the procedure. As prior studies in medical imaging have shown,^21,22^ if the transfer learning model overfits to this small, specific window, this may potentially make it worse at prediction compared to the global RF model.

On the other hand, our calibration analysis suggests that there may be some benefit of tuning beyond the first few minutes of surgery. This was indicated by the dramatic decrease in number of skipped LOSO folds with increasing adaptation windows, indicating increased capture of clinically relevant information.

### The unbiased judge: harnessing ensemble methods to reveal insights into medical machine learning

Ensemble methods are commonly used to mix behaviors of various models to boost the accuracy for more robust predictions.^23,24^ They can also be used to reveal insights into methodologies and data.

One of the first questions we explored is the difference in performance of the ensembled RF-TL with various combination strategies and RF tree sizes. While there was no difference in performance between 50 or 200 trees, a one-layer meta-learner produced a significant difference favoring the 200-tree RF-TL. This is indicative of the increased stability (lower variance) of increasing tree size, which smooths decision boundaries at the cost of increasing computation burden and model size linearly.^25^

The performance of the 200-tree RF-TL was further improved by the employment of a two-layer meta-learner. Consistent with earlier conclusions, this suggests that prediction mechanism using the engineered data is non-linear, which is suited well for RF’s robust predictive power.

This conclusion is further supported by the GateNet’s arbitrating behavior. GateNet’s top permutation importance features closely mirror that of the Random Forest model, indicating high trust. The alpha analysis suggests that in most cases, the best strategy to minimize error is to trust RF over TL, most likely due to the reasons outlined earlier in the discussion.

The clinical implications are substantial. Our findings suggests that for the purposes of nociceptive signal detection, a less computationally intensive, more interpretable, and easier-to-deploy Random Forest model is a superior approach. Especially as interpretability and scalability is at the forefront of discussion in every medical ML/AI tool, pursuing complex, “black box” models like TCNs may not be the best path forward. Instead, proper feature engineering and simple, interpretable models such as Random Forest may offer superior scalability and increase clinician trust without sacrificing accuracy.

### Final remarks

In this comprehensive evaluation of models for nociception detection, we found that a robust, supervised Random Forest model trained on engineered physiological features established a high-performance benchmark. While deep transfer learning offers a promising paradigm for patient-specific adaptation, our results indicate it provided no significant performance gain in this setting. Furthermore, we demonstrate the utility of ensemble methods such as a gated ensemble network as a diagnostic tool, which automatically determined the marginal value of the transfer learning component to be negligible. These findings underscore the critical importance of benchmarking against strong classical models and suggest that for this clinical application, a simpler, more efficient model may be the better solution.

## Data Availability

All data was sourced from Subramanian et al. on PhysioNet under data usage agreement and proper citations in the manuscript. All code and analysis can be provided upon reasonable request. The authors plan to upload their code on GitHub.

https://physionet.org/content/multimodal-surgery-anesthesia/1.0/

